# Clinical Determinants and Predictors for Prognosis of SARS-CoV-2 Infected Pediatric Patients in Saudi Arabia

**DOI:** 10.1101/2021.07.28.21261284

**Authors:** Khalid Mohamed Adam, Bahaeldin K. Elamin, Jaber A. Alfaifi, Mohammed Abbas

**Affiliations:** College of Applied Medical Science, University of Bisha, Bisha 61922. Saudi Arabia; Department of basic Medical Sciences, College of Medicine, University of Bisha, Bisha 61922, Saudi Arabia; Department of Child health, College of Medicine, University of Bisha, Bisha 61922, Saudi Arabia; Pediatric department, College of Medicine, Arabian Gulf University, Bahrain

**Keywords:** COVID-19, Pediatric, Children, Severe, Saudi Arabia, Comorbidity

## Abstract

**Background:** The significant variations in clinical characteristics and outcomes of COVID-19 that range from asymptomatic to severe fatal illness entail searching for potential prognostic determinants to help predict the disease course and early detection of patients at risk of developing life-threatening complications. Although children are less commonly infected by SARS-CoV-2 than their adult counterparts, and their symptoms are generally milder, a severe type of COVID-19 cannot be precluded.

**Methods:** At first, demographic, clinical, laboratory measurement data, and outcomes for 26 COVID-19 infected children of less than 12 years of age, admitted to King Abdallah Hospital, Bisha, Saudi Arabia, were retrieved from the electronic medical records for the observational retrospective study.

Later, electronic and manual database searches were carried out for pediatric severe COVID-19-related articles. The relevant data from 20 eligible studies and the present retrospective study were analyzed to assess the association of demographic characteristics and comorbidities with COVID-19 severity.

**Results:** In the retrospective study, 5 (19%) of the children presented with severe symptoms admitted to PICU, 18 (69%) presented with cough, 5 (19%) with diarrhea, 7 (27%) with underlying comorbidities, 4 (15%) with respiratory illnesses, 3 (12%) with cardiovascular diseases and 2 (8%) were obese. None of the patient characteristics showed any significant association with COVID-19 severity.

Of the 21 studies selected for meta-analyses, 14 studies were included in the analysis of the association between any comorbidity and disease severity, resulting in OR: 2.69, 95%CI: 1.38 – 5.26, P < 0.05, for analysis of the association between cardiovascular comorbidities and disease severity 14 studies were included giving OR: 4.06, 95%CI: 1.86 – 8.87, P < 0.05, for analysis of the association between respiratory comorbidity and disease severity 15 studies were included giving OR: 2.05, 95%CI: 1.54 – 2.74, P < 0.05, for analysis of the association between obesity and disease severity 10 studies were included, giving OR: 2.48, 95%CI: 1.16 – 5.32, P < 0.05, for analysis of the association between age <10 years old and diseases severity, 16 studies were included, giving OR: 0.80, 95%CI: 0.65 – 0.97, P < 0.05, and for analysis of the association between female gender and disease severity, 19 studies were included, giving OR: 0.83, 95%CI: 0.59 – 1.18, P > 0.05.

**Conclusion:** It can be concluded that COVID-19 pediatric patients with underlying comorbidities, being cardiovascular, respiratory, or obesity, are at high risk of developing severe illness, and young age has a protective role against the disease severity.

## Introduction

The fall of 2019 has marked the emergence of acute respiratory tract infection of unknown etiology in Wuhan, China, the cause of which was later discovered to be a large enveloped, positive-sense single-stranded RNA virus that belongs to the Betacoronavirus genus group 2, of the family Coronaviridae [1]. It was designated as severe acute respiratory syndrome Coronavirus type 2 (SARS-COV-2) and the disease as Coronavirus disease 2019 (COVID-19). This disease, along with the severe acute respiratory syndrome virus (SARS) and respiratory syndrome of the Middle East (MERS), formed three major outbreaks of zoonotic origin in the last two decades with devastating impact on the global healthcare system, economy, and almost all walks of life [2].

In March 2020, the World Health Organization declared the disease a global pandemic with exponentially increasing infection rates since its identification. The virus is primarily transmitted via respiratory droplets, but other routes such as direct contact with contaminated surfaces and the fecal-oral route were reported [3,4]. The infectivity and pathogenesis of SARS-Cov-2 are determined mainly by the route of transmission and host cell entry [5,6]. The receptor-binding domain (RBD) of the viral spike protein mediates viral entry by binding to angiotensin-converting enzyme 2 (ACE-2) on the host cell surface (cell 18). For efficient host cell entry, the viral spike protein is activated by the proteolytic action of TMPRSS2 and lysosomal protease cathepsins [7]. COVID-19 is associated with widely variable clinical presentations, ranging from asymptomatic to severe [8]. The disease also affects different body organs and systems differently. The respiratory system is the first to be affected, where the injury of lung endothelial and alveolar cells and vascular myocytes due to viral multiplication results in exudate accumulation, hyaline membrane formation, and fibrosis [9,10] with a characteristic inflammatory reaction and vascular angiogenesis [11,12]. The viral infection can also lead to ischemic heart disease through thrombosis and plaque rupture caused by the cytokine storm, inflammation, hypercoagulability, and platelet activation [13-16], the early manifestation of nervous system infection is the loss of smell, and this may progress into confusion and delirium in some cases [17]. Although angiotensin-converting enzymes -2 receptors (ACE2) are highly expressed in the cells of the gastrointestinal tract (GIT), few patients show GIT symptoms such as diarrhea, nausea and, loss of appetite, and some patients show alterations in liver function and bile duct injury [17]. The viral infection and the consequential cytokine storm were reported to have deleterious effects on renal functions, eyes, skin, and different physiological processes [18-21].

In Saudi Arabia, the number of total cases during the last year of the pandemic showed a steady increase, starting from a few cases in February 2020 to over 380000 cases in March 2021, with 6570 deaths, while the number of active cases fluctuated dramatically, reaching a peak of over 60000 during June - August 2020 and then the number declined to reach over 3000 cases in March 2021. Similarly, the number of critical cases peaked in July 2020, bordering 2300 cases, and declined to reach a little over 500 in March 2021 [22]. To the best of our knowledge, the prognostic indicators associated with disease outcomes and severity in pediatrics have not been investigated in the Saudi population. This study aimed to unravel the role of different demographic, laboratory, and clinical characteristics as prognostic indicators for COVID-19 severity in pediatric patients.

## Materials and Methods

### Study design and setting

In this retrospective, observational study, that ethically and technically approved by the institutional review board of deanship of scientific research, University of Bisha, the clinical, demographic, and comorbidity data on presentation, treatment, and outcomes of all children aged less than 12 years admitted to King Abdalla Hospital, Bisha, Saudi Arabia, between March 1, 2020, and April 28, 2021, with a laboratory-confirmed SARS-CoV-2 infection, retrieved from the electronic medical record (EMR). Severe disease was defined as requiring admission to a pediatric intensive care unit (PICU)

### Meta-analysis

#### Literature search strategy

The systematic literature search, on the other hand, was conducted following the Preferred Reporting Items for Systematic Reviews and Meta-analysis (PRISMA) and the Meta-analyses Of Observational Studies in Epidemiology (MOOSE) guidelines [23,24]. Medline, Embase, Web of Science, and ProQuest databases were queried for records published from January 1, 2020, until May 1, 2021, using the combination of the following terms: “COVID-19”, “SARS-CoV-2”, “novel Coronavirus,” “2019-nCoV”, “Sever*,” “Comorbidit*,” “Clinical characteristics,” “Clinical features,” to look for studies in which investigated samples were divided into age groups. Later, the search terms “Pediatr*,” “child*,” “infant,” “newborn” were included in the search. BioRxiv, MedRxi, Open Grey, and Open Science Framework (OSF) preprints were queried for grey literature. In addition, the reference lists for all potentially eligible articles were manually searched.

All records were initially screened, and duplicates were removed.

#### Study eligibility criteria and quality assessment

Eligible studies included in the quantitative synthesis met the following inclusion criteria: 1) cross-sectional cohort design, 2) studies explicitly mentioning mild, moderate, and severe states of the patients studied, 3) studies including pediatrics or age group of <12 years, 4) studies reported data on comorbidities, 5) studies reported the odds ratio and corresponding 95% confidence interval or enough data to calculate these values, 6) published in English language only.

Studies excluded were adults-only studies or studies where pediatric data were indistinguishable from adult data, studies reported no severe cases, reviews, editorials, case studies, and case series.

Two reviewers independently assessed the quality of the included studies using the Newcastle Ottawa Scale (NOS) for cross-sectional studies. We assessed criteria that cover selection, comparability, and exposure. Studies were awarded a maximum of one point for each item in the selection and exposure categories and a maximum of two points for comparability, with a total of 9 attainable points. Studies with a score *≥ 7* regarded high quality, 5 – 7 moderate quality, and a score <5 poor quality.

The scoring was carried out by two reviewers independently, and disagreement was resolved by consensus.

#### Data extraction

Data from eligible studies were extracted by two reviewers independently and vetted by two different reviewers using a data extraction table. The extracted data included the name of the first author, year of publication, population studied, number of severe and moderate/mild cases according to age, gender, with any comorbidity with respiratory comorbidities, with cardiac comorbidities, with obesity, and elements of bias risk assessment.

#### Statistical analyses

Patient demographics, clinical characteristics, and laboratory findings were summarized using median, minimum, and maximum values, or frequency and percentage where appropriate. The outcomes were compared using the Wilcoxon rank sum test, while logistic regression was used to test the association between potential risk factors and disease severity. Results were considered statistically significant at *P* < 0.05 using SPSS software (IBM SPSS Statistics for Windows, Version 25.0. Armonk, NY: IBM Corp.).

The meta-analysis was performed using Prometa3 software. We computed the pooled odds ratio (OR) and the corresponding 95% confidence interval (CI) using the random-effects model (DerSimonian Laird method). The heterogeneity among included studies was estimated using Cochran Q statistic and *I*^2^. Heterogeneity was considered insignificant with *I*^2^ < 40%, moderate heterogeneity with *I*^2^ between 40% - 60%, and substantial heterogeneity with *I*^2^ > 60%. Publication bias was evaluated with the funnel plot, Egger’s linear regression test, and Begg’s rank correlation test, with *P*. value < 0.1 indicating potential bias. The sensitivity analysis was also carried out to assess the influence of each individual study.

## Results

Twenty-six children hospitalized with COVID-19 during the study period were included, constituted of 14 (54%) females, 11(42%) less than a year, 13 (50%) from 1 – 5 years, and 2 (8%) more than five years of age. The majority of the pediatric patients, 21 (81%) admitted to the regular care unit, whereas 5 (19%) suffered a severe illness that required PICU admission (Table 1). On admission, 18 (69%) presented with cough, 5 (19%) with diarrhea, and 18 (69%) with chest distress. Seven children (27%) were known to have preexisting comorbidities; the most common comorbidities were respiratory illnesses 4 (15%) and cardiac 3 (12%). Two children (8%) were obese.

**Table 1:** Demographic and clinical characteristics of COVID-19 patients

| Variable | COVID-19 patients<br>(n) % |
| --- | --- |
| **Age:** |  |
| <1 year | 11 (42%) |
| 1-5 years | 13 (50%) |
| >5 years | 2 (8%) |
| **Gender:** |  |
| Male | 12 (46%) |
| Female | 14 (54%) |
| **Cough:** |  |
| Yes | 18 (69%) |
| No | 8 (31%) |
| **Diarrhea:** |  |
| Yes | 5 (19%) |
| No | 21 (81%) |
| **Underlying comorbidity** |  |
| Yes | 7 (27%) |
| No | 19 (73%) |
| **Respiratory comorbidity** |  |
| Yes | 4 (15%) |
| No | 22 (85%) |
| **Cardiovascular comorbidity** |  |
| Yes | 3 (12%) |
| No | 23 (88%) |
| **Obesity** |  |
| Yes | 2 (8%) |
| No | 24 (92%) |
| **Disease severity** |  |
| Yes | 5 (19%) |
| No | 21 (81%) |

Table 2 shows the laboratory findings in patients with severe illness in comparison with those presented with mild or moderate type of disease. The two groups showed lack of statistically significant difference in WBC (*P* > 0.05), neutrophils (*P* = 0.974), lymphocytes (P > 0.05), hemoglobin (*P* > 0.05), platelets (*P* > 0.05), CRP (*P* > 0.05), ALT (*P* > 0.05), AST (*P* > 0.05), albumin (*P* > 0.05), serum creatinine (*P* > 0.05), Na (*P* > 0.05), K (*P* > 0.05), Cl (*P* > 0.05). Similarly, the odds ratio and 95% confidence interval values showed no association between disease severity and patient characteristics (Table 3).

**Table 2:** Laboratory parameters and comparison between severe and nonsevere patients

| Variable | Overall COVID -19 patients results |  |  | Severe Vs. Non-severe COVID-19 patients |
| --- | --- | --- | --- | --- |
|  | Median | Mini | Max | *P. value* |
| WBC ($\times 10^3/\mu\text{l}$) | 9.5 | 2.3 | 23 | 0.345 |
| Neutrophils (%) | 39 | 2 | 90 | 0.974 |
| Lymphocytes (%) | 42 | 3.7 | 87 | 0.204 |
| Hb (g/dL) | 11 | 6.8 | 13.7 | 0.947 |
| Platelet ($\times 10^3/\mu\text{l}$) | 331 | 26 | 580 | 0.922 |
| CRP (mg/L) | 25 | 10 | 48 | 0.064 |
| ALT (U/L) | 31 | 5 | 142 | 0.580 |
| AST (U/L) | 39 | 8.3 | 76 | 0.356 |
| Albumin (g/L) | 36 | 28 | 48 | 0.944 |
| S. Creatinine (mg/dl) | 0.33 | 0.19 | 0.80 | 0.297 |
| Na (mEq/L) | 137 | 131 | 146 | 0.203 |
| K (mEq/L) | 4.45 | 4 | 6 | 0.744 |
| Cl (mEq/L) | 103 | 97 | 111 | 0.793 |

**Table 3:** Association of patient characteristics and COVID-19 severity

| **Variables** | **OR (95% CI)** | **P. value** |
| --- | --- | --- |
| **Demographic data** |  |  |
| Age <10 years | 0.885 (0.165 – 4.746) | 0.887 |
| Gender | 1.311 (0.168 – 10.211) | 0.796 |
| **Symptoms** |  |  |
| Cough | 1.429 (0.173 – 11.806) | 0.740 |
| Chest distress | 1.667 (0.220 – 12.617) | 0.621 |
| Diarrhea | 0.250 (0.029 – 2.183) | 0.210 |
| **Laboratory measures** |  |  |
| WBC | 1.076 (0.876 – 1.321) | 0.485 |
| Neutrophils | 1.006 (0.966 – 1.049) | 0.764 |
| Lymphocytes | 0.965 (0.917 – 1.015) | 0.170 |
| Hemoglobin | 0.878 (0.492 – 1.567) | 0.660 |
| platelets | 1.001 (0.993 – 1.009) | 0.862 |
| CRP | 0.924 (0.820 – 1.040) | 0.188 |
| ALT | 1.034 (0.981 – 1.091) | 0.215 |
| AST | 0.947 (0.869 – 1.031) | 0.208 |
| Albumin | 1.069 (0.834 – 1.371) | 0.598 |
| Serum Creatinine | 1.034 (0.969 – 1.103) | 0.319 |
| Na | 1.161 (0.906 – 1.487) | 0.238 |
| K | 0.645 (0.90 – 4.612) | 0.662 |
| Cl | 0.997 (0.776 – 1.281) | 0.980 |
| **Comorbidity** |  |  |
| Any comorbidity | 6.38 (0.78 – 51.78) | 0.083 |
| Respiratory | 1.50 (0.12 – 18.44) | 0.751 |
| Cardiovascular | 13.33 (0.91 – 196.37) | 0.059 |
| Obesity | 0.71 (0.03 – 17.06) | 0.832 |

The systematic search for relevant literature in Medline, Embase, Web of Science, and ProQuest databases resulted in the retrieval of 767 articles, in addition to 37 articles identified through grey literature and manual search. Following the removal of duplicates and irrelevant articles, 329 articles were subjected to title and abstract screening, which resulted in the exclusion of 147 articles. For the remaining 155 articles, a thorough full-text evaluation was conducted, leading to further exclusion of 135 articles due to lack of extractable data; the pediatric age group included patients up to 20 years old, indistinguishable pediatric data, and different study designs. Ultimately, 20 articles were included in this meta-analysis along with the present retrospective study (Fig. 1). The 21 included studies did not cover the variables to be investigated in this meta-analysis; hence, the meta-analyses conducted for the association of disease severity with any comorbidities included 14 studies with a sample size of 336363 patients, with age less than ten years included 16 studies with a sample size of 45807 patients, with female gender, included 19 studies with a sample size of 35271 patients, with respiratory comorbidities included 15 studies with a sample size of 14427 patients, with cardiac comorbidities, included 14 studies with a sample size of 14810 patients, and with obesity included ten studies with a sample size of 1730 patients.

**Fig. 1:**
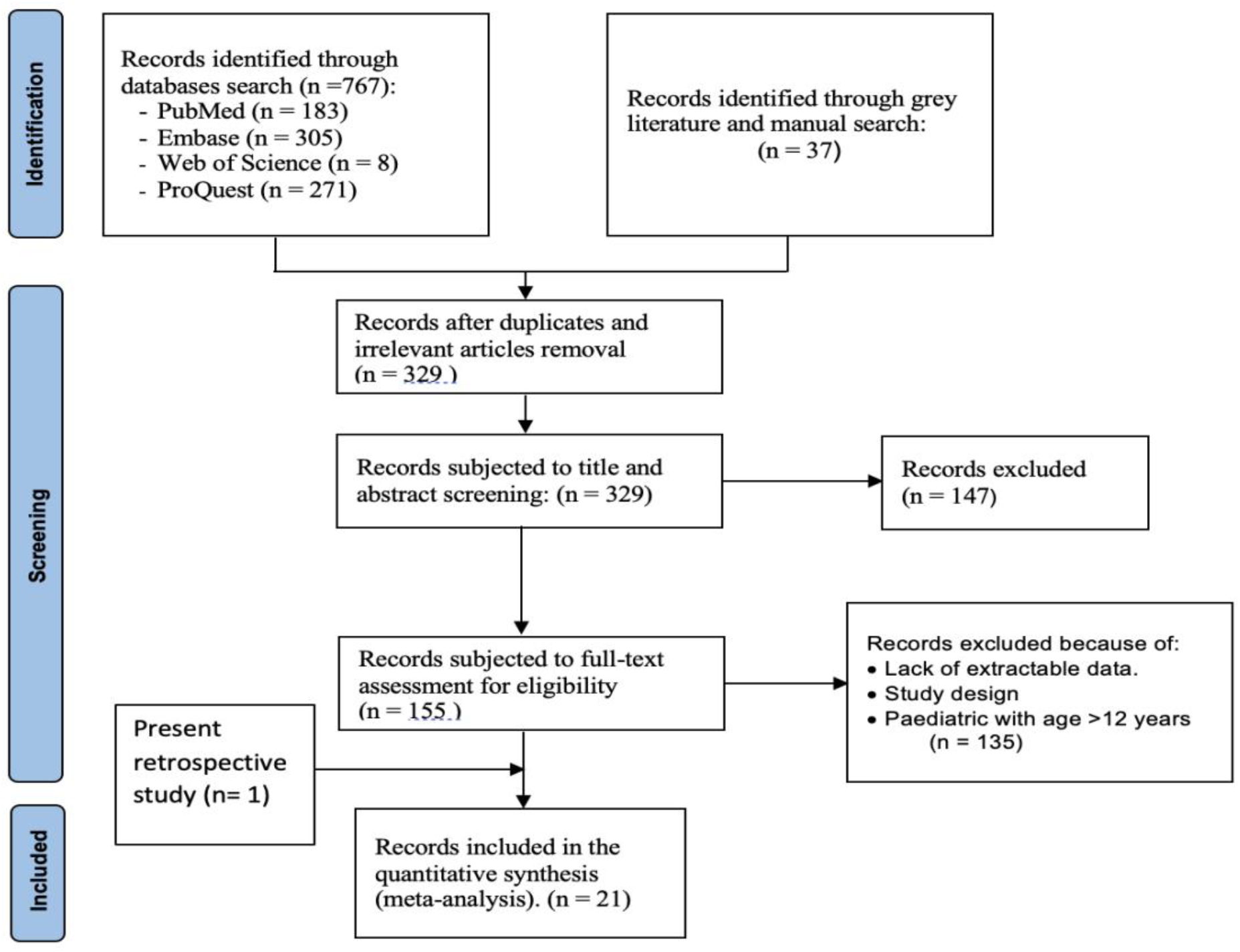
PRISMA flowchart of studies included in the quantitative synthesis

These 21 studies constituted of 9 studies from the United States, two studies each from Saudi Arabia and Brazil, and one study each from Italy, China, Oman, Europe, Spain, England, Romania, Thailand.

We estimated the pooled effect size and the level of heterogeneity among the studies used to determine association between disease severity and study variables, namely, presence of any comorbidity (OR: 2.69, 95%CI: 1.38 – 5.26, *P* < 0.05) (Q = 131.50, *P*. value < 0.05, *I*^2^ = 90.11%), age less than 10 years (OR: 0.80, 95%CI: 0.65 – 0.97, *P* < 0.05) (Q = 24.96, *P*. value = 0.050, *I*^2^ = 39.91%), female gender (OR: 0.83, 95%CI: 0.59 – 1.18, *P* > 0.05) (Q = 54.64, *P*. value < 0.05, *I*^2^ = 67.06%), respiratory comorbidities (OR: 2.05, 95%CI: 1.54 – 2.74, *P* < 0.05) (Q = 15.51, *P*. value > 0.05, *I*^2^ = 9.71%), cardiac comorbidities (OR: 4.06, 95%CI: 1.86 – 8.87, *P* < 0.05) (Q = 71.40, *P*. value < 0.05, *I*^2^ = 81.79%), and obesity (OR: 2.48, 95%CI: 1.16 – 5.32, *P* < 0.05) (Q = 17.27, *P*. value < 0.05, *I*^2^ = 47.88%) (Fig. 2 – 7).

**Fig. 2:**
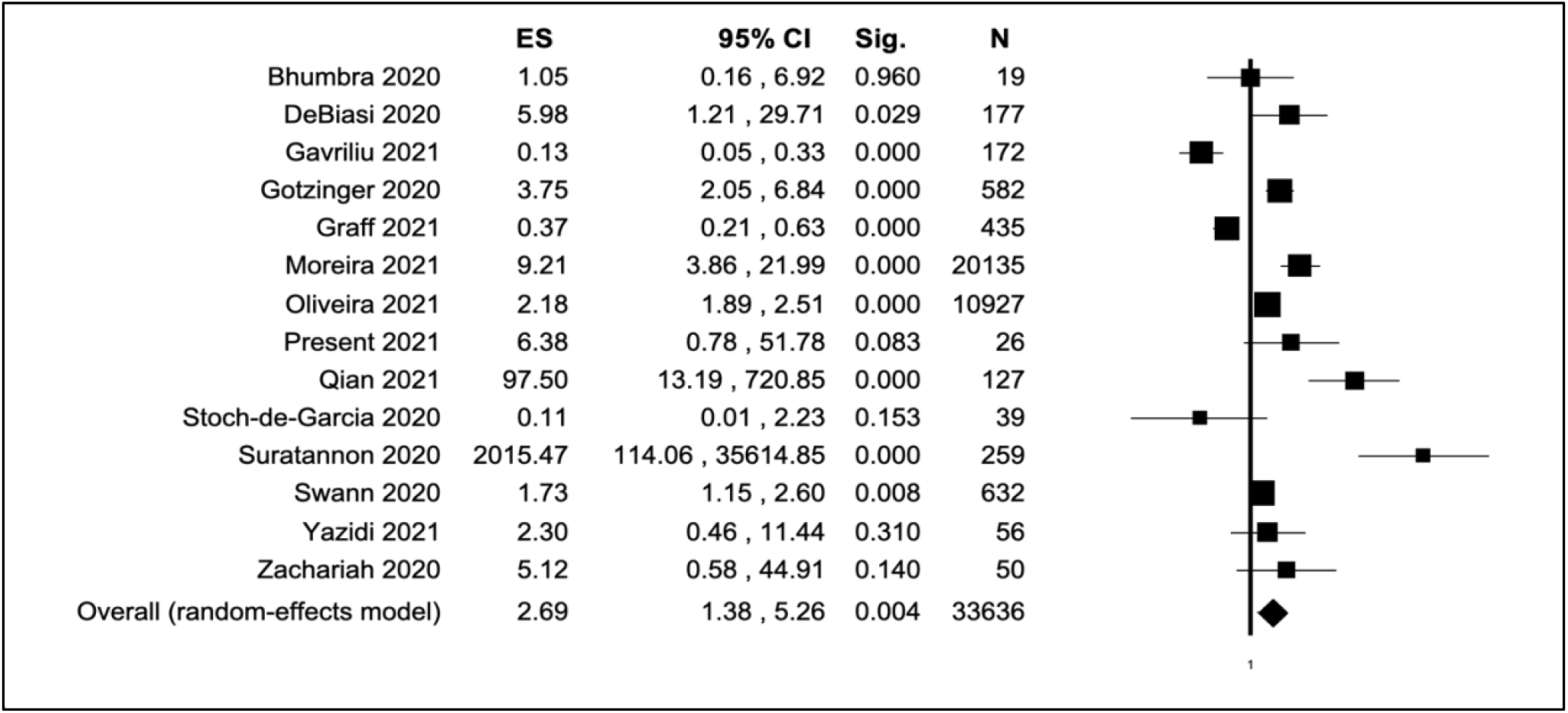
Forest plot of the association between underlying comorbidity and COVID-19 severity

**Fig. 3:**
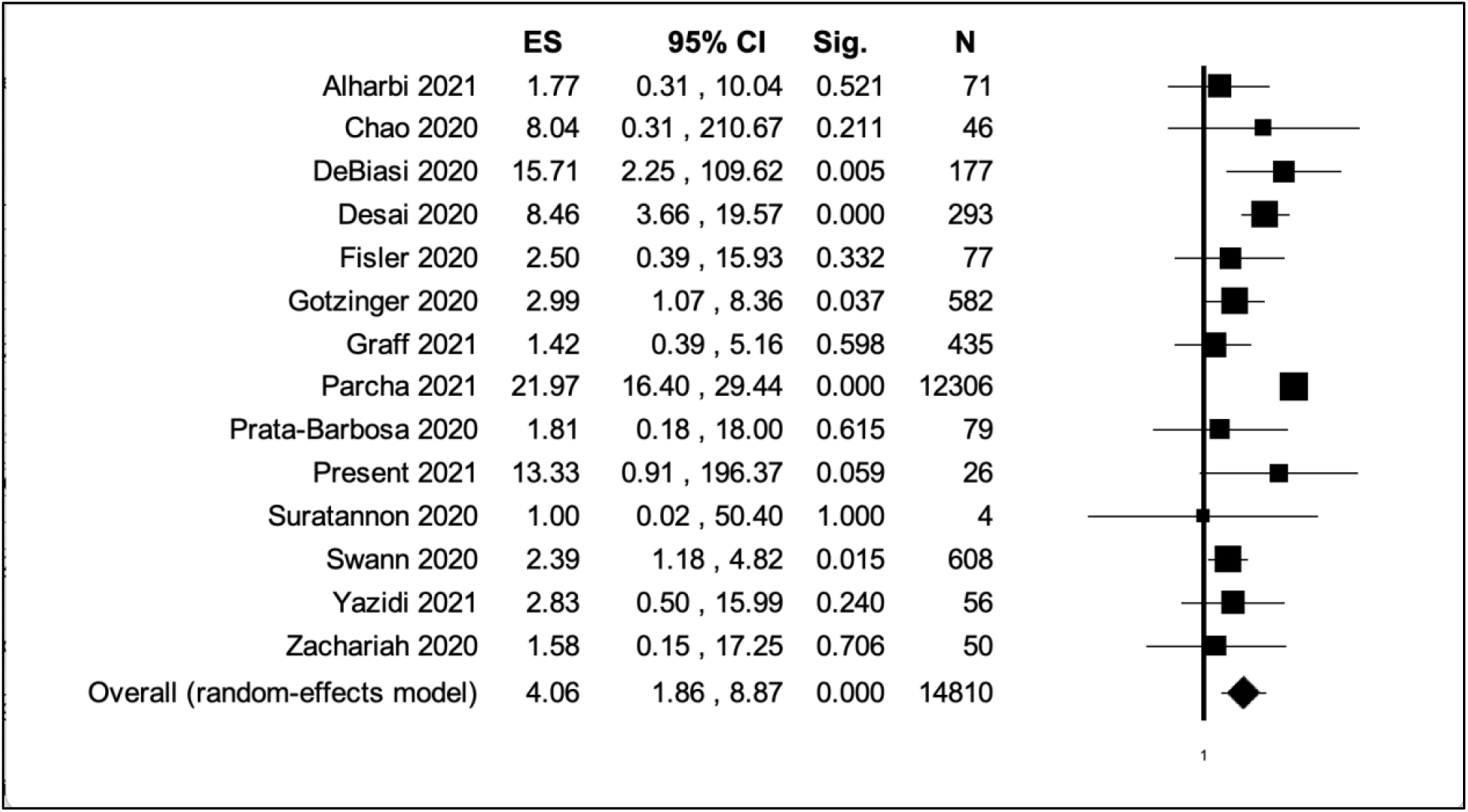
Forest plot of the association between comorbid CVD and COVID-19 severity

**Fig. 4:**
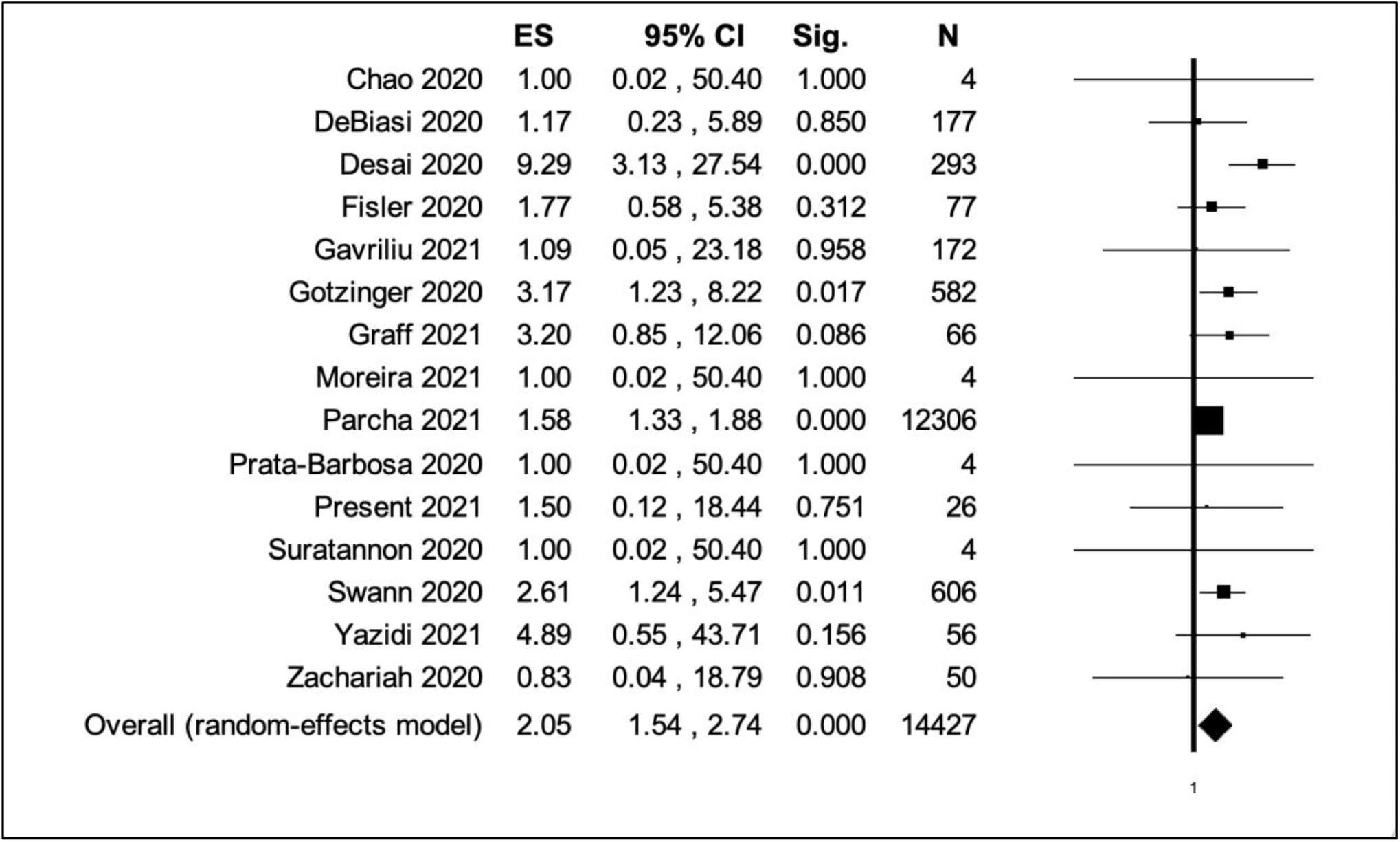
Forest plot of the association between comorbid respiratory diseases and COVID-19 severity

**Fig. 5:**
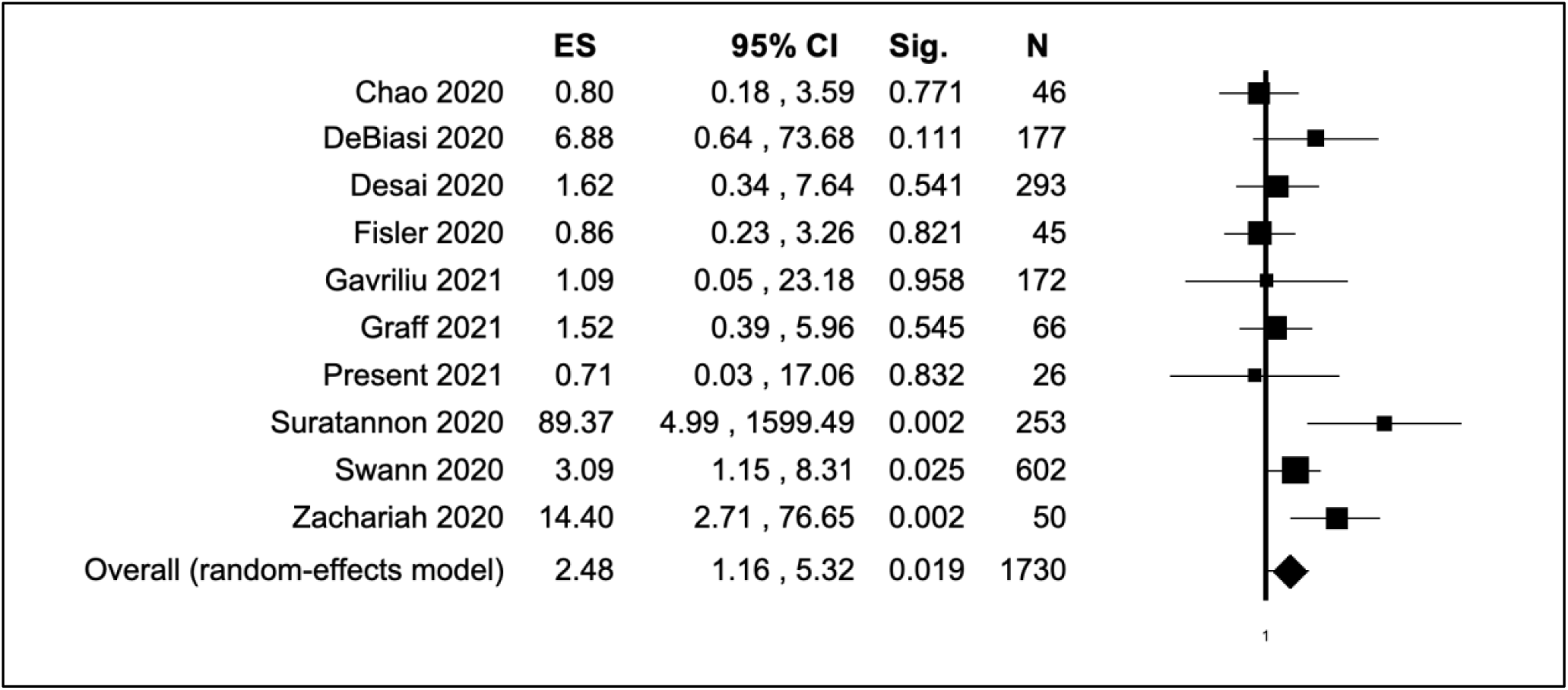
Forest plot of the association between obesity and COVID-19 severity

**Fig. 6:**
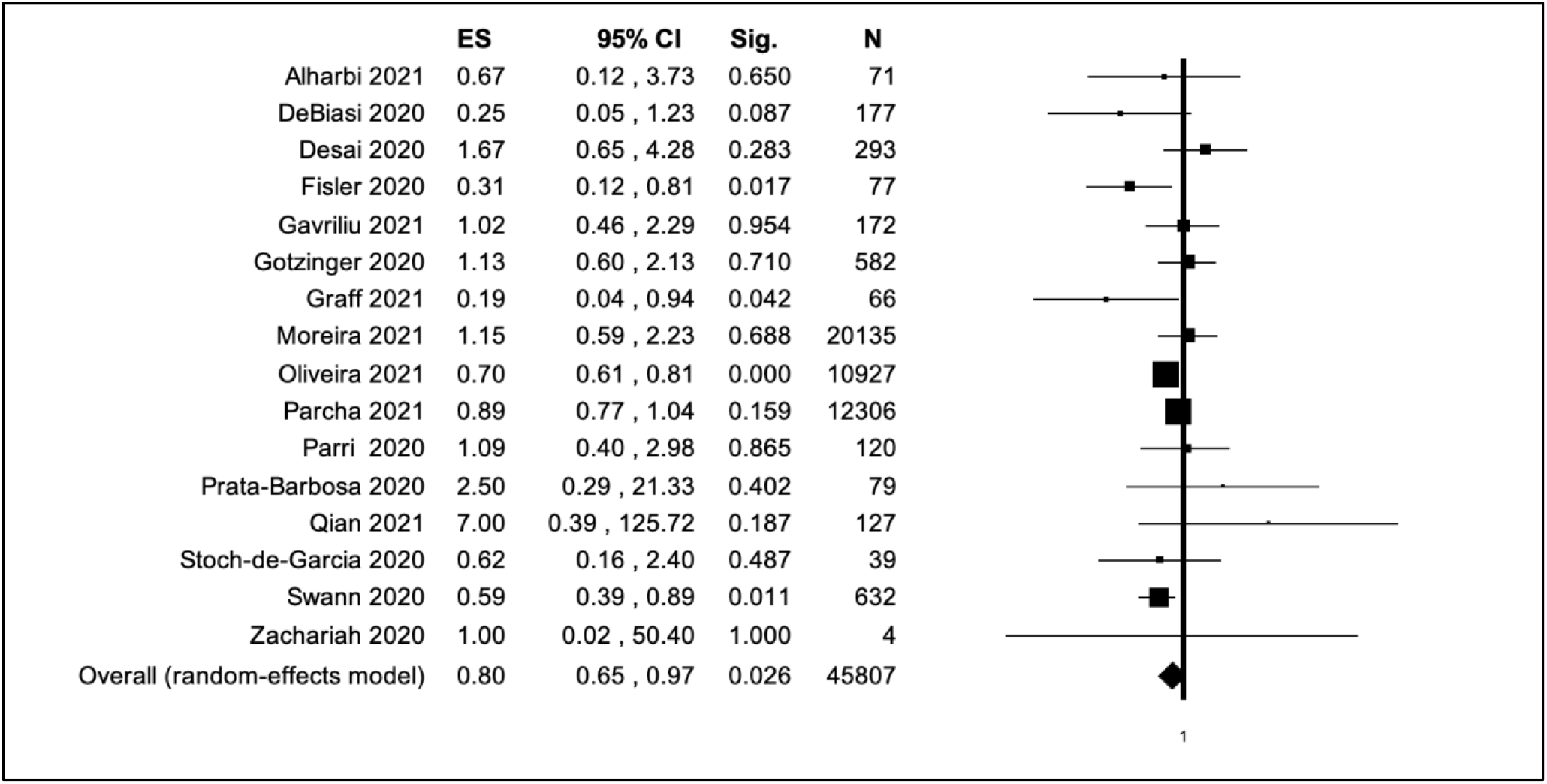
Forest plot of the association between age <10 years and COVID-19 severity

**Fig. 7:**
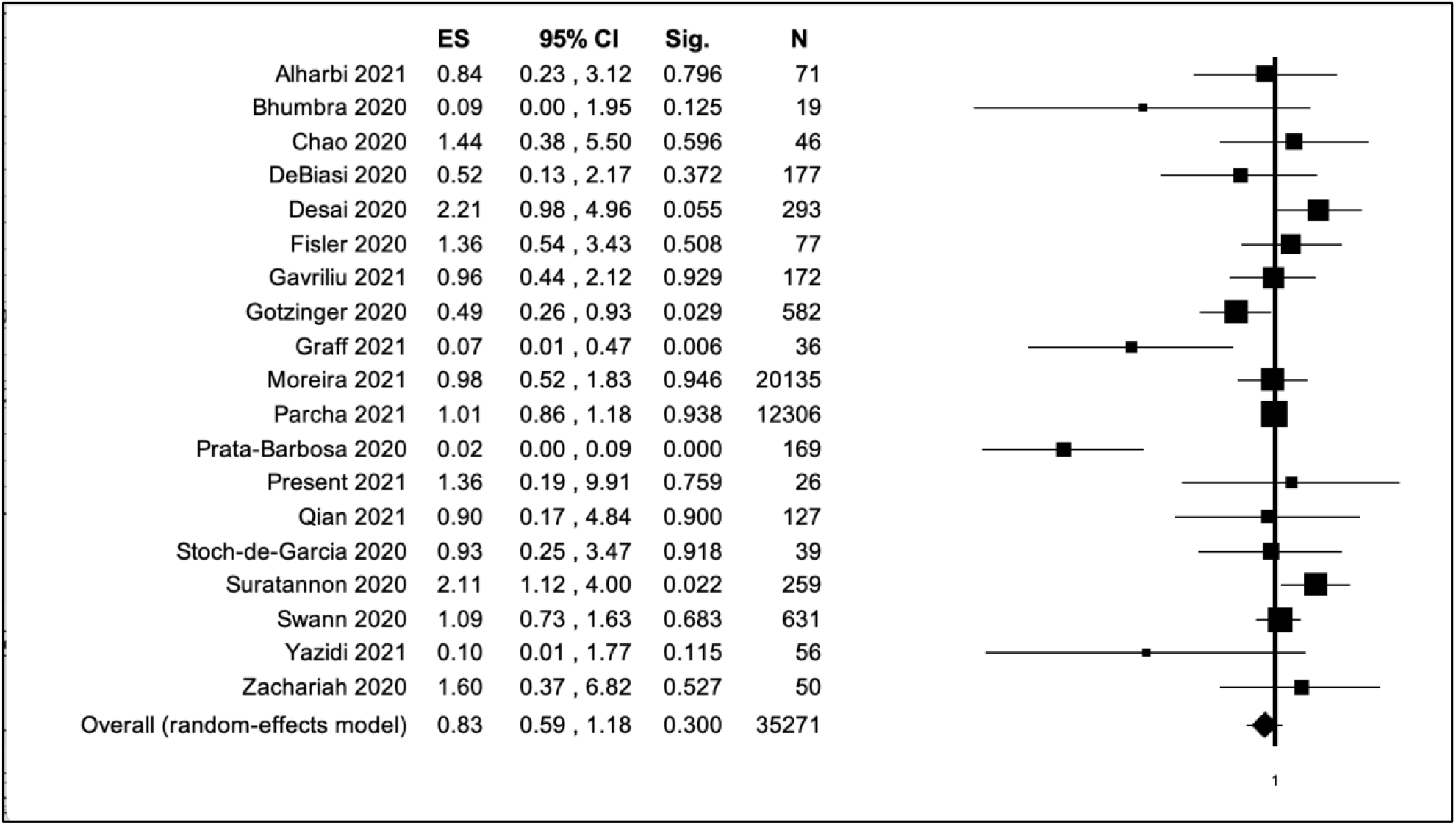
Forest plot of the association between female gender and COVID-19 severity

### Sensitivity analysis and publication bias

The sensitivity analysis was carried out by removing one study at a time during the analysis. The lack of significant change in the pooled effect size for studies of both viruses indicated stability of the present meta-analyses. The publication bias was assessed with Begg’s rank correlation test, Egger’s linear regression test, and funnel plot. None of the meta-analyses showed publication bias. The results of meta-analysis of disease severity and any underlying comorbidities; Begg’s test (*Z* = 0.38, *P* > 0.05), Egger’s test (t = 0.34, *P* > 0.05) (Fig. 8A), the meta-analysis of disease severity and age of <10 years; Begg’s test (*Z* = -0.18, *P* > 0.05), Egger’s test (t = 0.25, *P* > 0.05) (Fig. 8B), the meta-analysis of disease severity and female gender; Begg’s test (*Z* = -1.99, *P* < 0.05), Egger’s test (t = -1.36, *P* > 0.05) (Fig. 8C), the meta-analysis of disease severity and respiratory comorbidities; Begg’s test (*Z* = -1.34, *P* > 0.05), Egger’s test (t = 1.14, *P* > 0.05) (Fig. 8D), the meta-analysis of disease severity and cardiac comorbidities; Begg’s test (*Z* = 1.15, *P* > 0.05), Egger’s test (t = -3.31, *P* < 0.05 (Fig. 8E), and the meta-analysis of disease severity and obesity; Begg’s test (*Z* = 0.80, *P* > 0.05), Egger’s test (t = 0.66, *P* > 0.05) (Fig. 8F).

**Fig. 8:**
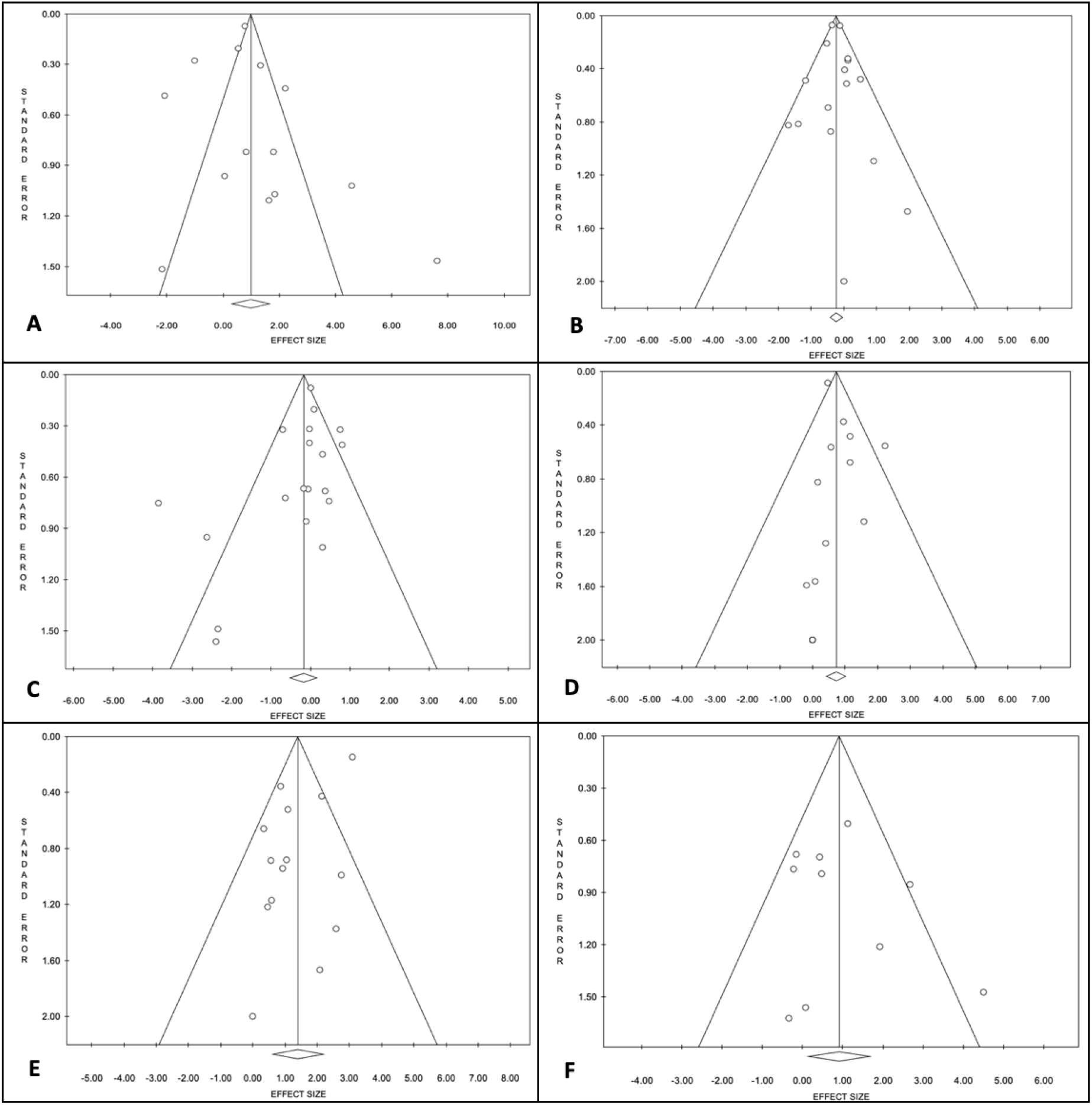
Funnel plot for estimation of publication bias, (**A**) disease severity and underlying comorbidity, (**B**) disease severity and age, (**C**) disease severity and female gender, (**D**) disease severity and respiratory comorbidity, (**E**) disease severity and CVD comorbidity, (**F**) disease severity and obesity

## Discussion

In the present study, we attempted to explore the association of the demographic, clinical, and laboratory data of COVID-19 pediatric patients with the severity of the disease, the indicator of which was PICU admission [25]. Initially, the contingency of disease severity on patient characteristics was explored in a cohort of 26 hospitalized pediatric patients, and the results were then included in a meta-analysis that encompassed 21 studies. In our retrospective cohort, a small proportion of the patients, 19%, required PICU, while the rest, although hospitalized, but presented with mild to moderate symptoms, which goes in concordance with reports by Bellino S. *et al*.. [26] in Italy, Hernandez-Garduno E. *et al*.. [27] in Mexico, and Du H. *et al*.. [28] in China, suggesting that young children are less prone to severe type of COVID-19. The small number of severely ill pediatric patients in the present study could be the reason that neither of the variables studied showed any statistically significant association with disease severity; conversely, multiple studies reported an association of disease severity with comorbidity [29,26,30,31], age [32,33], obesity [34,26], CRP [33,28]. The discrepancy between the present study’s findings and previously published reports [25,35] was also noted in the association of disease severity with laboratory measurements, where both patients with severe and nonsevere COVID-19 in this study showed readings within the normal range for all parameters studied.

These findings were then combined with that of other similar studies in a meta-analysis to unravel the association of different factors with the severity of COVID-19.

The meta-analysis result of the association between disease severity and the presence of underlying comorbidity suggests that the risk of developing severe COVID-19 is 2.69-folds higher in children with any underlying comorbidity, a finding that is corroborated by previous reports on both children and adults [36-38]. A similar statistically significant increase in the risk of severe COVID-19 has been noted with chronic respiratory comorbidity by 2.05-fold, cardiovascular comorbidity by 4.06-fold, and obesity by 2.48-fold, but not female gender. While age less than 10 years is a protective factor. Comorbid cardiovascular diseases had noticeably the strongest association with disease severity, followed by obesity and respiratory diseases. Although the nature of the association between cardiovascular diseases and the severity of COVID-19 is not well defined, the expression of ACE-2 receptors on the pericytes and muscle cells of the heart and overexpression in cases of heart disease [39] as well as the increased secretion of inflammatory cytokines, and compromised immune system [40] can be suggestive of the mechanisms behind this association.

The poor prognosis of COVID-19 associated with obesity was reported by several pediatric and adult studies [38,41]. Obesity is believed to promote the production of inflammatory cytokines, tumor necrosis factor *α*, and pro-inflammatory macrophages [38], in addition to the adversary effect on important host immune response players such as natural killer (NK) cells and mucosal-associated invariant T cells (MAIT) [42]. Moreover, the role of respiratory tract diseases in potentiating severe COVID-19 can be attributed to the overexpression of ACE-2 receptors on lung cells, increased mucous production, and compromised immunity [40]. In contrast to studies that reported more severe COVID-19 in children younger than five years old [43,44], we found no association between neither the child’s age nor gender with disease severity, which can be attributed to the significant heterogeneity across studies.

Although we rigorously analyzed data from many relevant COVID-19 studies, our study suffered some limitations, including heterogeneity across studies included for exploring the association of disease severity with any comorbidity, cardiovascular disease, and female gender but not age and respiratory tract diseases. This heterogeneity is believed to have affected the plausibility of some of our results. This study also focused retrospectively on specific comorbidities only, which entails more extensive prospective studies to establish the causality between severe COVID-19 and other comorbidities such as gastrointestinal, renal, genetic, and neurological diseases.

## Conclusion

In conclusion, this study described in part the lack of association between patient characteristics and the risk of severe COVID-19 in Saudi pediatric patients, and through meta-analysis it is inferred that age less than 10 years is a protective factor against disease severity while comorbidities of cardiovascular disease, respiratory diseases, and obesity are determinants of COVID-19 poor prognosis, and thus can be used to identify patients require close attention.

## Data Availability

Data are available upon reasonable request from the authors

## Data Availability

The datasets used and/or analyzed during the current study are available from the corresponding author on reasonable request.

## Acknowledgment

We are indebted to the deanship of scientific research, University of Bisha, and the staff of King Abdalla Hospital, Bisha.

